# Polarization-Sensitive Optical Coherence Tomography Imaging of Posterior Staphyloma Edges in Eyes with High Myopia

**DOI:** 10.1101/2025.02.24.25322667

**Authors:** Yijin Wu, Hongshuang Lu, Changyu Chen, Jianping Xiong, Masahiro Yamanari, Hiroyuki Takahashi, Keigo Sugisawa, Michiaki Okamoto, Yining Wang, Ziye Wang, Tomonari Takahashi, Koju Kamoi, Kyoko Ohno-Matsui

## Abstract

**Purpose:** To investigate the arrangement and orientation of the collagen fibers in the inner and outer scleral layers at the edges of posterior staphylomas using polarization-sensitive optical coherence tomography (PS-OCT).

**Methods:** 195 eyes of 105 highly myopic patients who underwent PS-OCT examinations between August 2023 and May 2024 at Institute of Science Tokyo were studied. The PS-OCT images were analyzed to assess the orientation and birefringence of scleral collagen fibers at the staphyloma edges, covering macular and inferior staphylomas.

**Results:** Eighty-eight eyes of 60 highly myopic patients whose posterior staphylomas were confirmed in the ultra-widefield optical coherence tomography images underwent PS-OCT image processing. Thirty-six eyes from 27 patients were selected for further analysis. The optic axis images showed that 25 (18 upper and 7 lower) of 30 (83.3%) macular staphyloma edges had an aggregation of the inner scleral fibers, characterized by an arc-like gathering of the horizontal fibers along the edge. All eight upper edges from 8 eyes with an inferior staphyloma had an aggregation of the inner scleral fibers with relatively linear horizontal fibers across the macula. The birefringence images of the upper and lower edges of the macular and inferior staphylomas had different patterns, including a mixture of low and high, and relatively high birefringence.

**Conclusions:** An uneven aggregation of inner scleral fibers at the staphyloma edges in highly myopic eyes offers a more nuanced understanding of the pathogenesis of staphylomas. This finding also has pivotal implications for developing the targeted interventions to prevent the progression of staphylomas in eyes with high myopia.

## Introduction

A posterior staphyloma is a hallmark finding in eyes with pathologic myopia^1–3^. It is described as a circumscribed posterior outpouching of the ocular wall of the eye with a shorter radius of curvature than the surrounding regions.^4^ Highly myopic eyes with staphylomas mechanically stretch and damage the retina in the macular region and the optic nerve, resulting in visual loss^5,6^ and vision-threatening complications.^7,8^ Although the pathogenesis has not been conclusively determined, the sclera is regarded as the primary target tissue whose morphology and physiology play a key role in the development of posterior staphylomas, regardless of the initially affected tissue.

The sclera is a fibrous connective tissue that is mainly composed of collagen fibers, fibroblasts, and an amorphous ground substance.^9^ Scleral collagen fibers are crucially involved in eyeball shape maintenance and mechanical damage shielding^10^. Despite multiple studies, the exact pathogenesis of posterior staphylomas has yet to be elucidated. McBrien et al.^11^ examined cadaver eyes histologically and proposed that staphylomas in axially elongated eyes resulted from localized and progressive alterations in the orientation of the scleral collagen fiber bundles at the posterior pole. For inferior staphylomas, Maruko et al.^12^ used enhanced depth imaging optical coherence tomography (EDI-OCT) and noted a thickening of the upper edge of the sclera. However, conventional OCT lacks the ability to assess the organization of scleral collagen fibers and studies of OCT images focused on the thickness and curvature of the sclera.

Polarization-sensitive OCT (PS-OCT) is an advanced OCT technology offering improved functionality compared to traditional OCT^13^. PS-OCT generates contrast images based on the birefringence of the tissue,^14^ and it has been demonstrated a promising tool for *in vivo* imaging of the scleral collagen fibers in animals.^15,16^ Birefringence is an optical property of fibrous tissue in which the refractive indices that vary with the light’s polarization state.^17^ The slow axis, corresponding to the optic axis (OA) with a higher refractive index, aligns with the long axis of biological fibers. This property enables the determination of the orientation of the collagen fibers.

PS-OCT can measure both the magnitude of birefringence, which reflects the arrangement^18^ and density^19^ of the scleral collagen fibers, and their orientation by determining the orientation axis or the OA of the birefringence.^20,21^ PS-OCT-derived OA images can clearly differentiate the inner scleral layer, which runs radially from the optic nerve to the periphery; from the outer layer, which predominantly runs vertically in the posterior fundus.^19^ Ohno-Matsui et al.^19^ used PS-OCT to examine highly myopic eyes with dome-shaped macula (DSM), and observed that the DSM consisted of a thickened inner scleral layer that compressed the outer sclera without thickening it. Although DSMs and posterior staphylomas have different pathologies, both are manifested as inward scleral protrusions. Therefore, evaluating the sclera using PS-OCT may provide novel information on the pathogenesis of staphylomas. However, to the best of our knowledge, no studies have focused specifically on the scleral fibers at the posterior staphyloma edges and their distinct morphologic variations. Thus, the study aimed to investigate the arrangement and orientation of the collagen fibers in the inner and outer scleral layers at the staphyloma edges using PS-OCT.

## Methods

This case series study was conducted in accordance with the tenets of the Declaration of Helsinki. and the procedures were approved by the Ethics Committee of the Institute of Science Tokyo. We adhered to the reporting guideline for an uncontrolled case series^22^ to ensure that our findings are reported comprehensively and with transparency. All participants signed a written informed consent.

### Participants

PS-OCT images of 195 eyes of 105 patients with high myopia who underwent examinations between August 2023 and May 2024 at the Advanced Center of High Myopia were retrospectively analyzed. High myopia was defined as a refractive error (spherical equivalent) of < −6.00 diopters (D) or an axial length (AL) > 26.5 mm. The exclusion criteria were poor quality PS-OCT images caused by media opacities, including dense cataracts or vitreous opacities; prior vitreoretinal surgeries; and eyes with peripapillary posterior staphyloma because the staphylomatous areas were not fully covered by the macular-centered scan.

### Posterior Staphylomas

A posterior staphyloma was identified by detecting its edges in the ultra-widefield OCT (UWF-OCT) images as described by Shinohara et al.:^23^ (1) a gradual “thick-thin-thick” change in the choroidal thickness from the periphery to the edge of the staphyloma and then to the posterior pole; and (2) an inward protrusion of the sclera at the edge of the staphyloma. The images were acquired with the Xephilio OCT-S1 instrument (Canon, Japan) with a scan range of 23 mm × 20 mm. Because of the typically steeper nature of the upper or lower edges, we determined the staphyloma edges in the vertical section of the 12 radial OCT sections centered on the fovea. The types of the posterior staphylomas were categorized by the location and distribution of the outermost contour of the posterior staphyloma using the classification proposed by Ohno-Matsui et al.^3^ Based on the wide-field fundus photographs and UWF-OCT images, there were two main types of posterior staphylomas: (1) macular staphyloma that included the wide macular type (Curtin’s type I^2^) and the narrow macular type (Curtin’s type II^2^). In the wide macular staphylomas, the nasal edge extended beyond the nasal margin of the optic disc; and in the narrow macular staphylomas, the nasal edge was aligned with the nasal margin of the optic disc; (2) inferior staphyloma (Curtin’s type V^2^) that existed exclusively in the lower fundus, and its upper edge usually passed across the macular area.

### PS-OCT Examination and Image Processing

A prototype PS-OCT system (ROCTIA, Tomey Corporation, Japan) was used to evaluate the birefringent properties of the posterior segment of the eye. This system incorporates a polarization-sensitive optical interferometer and a swept laser with a central wavelength of 1050 nm and an A-scan rate of 100 kHz as described previously in detail.^24^ The retinal scans covered an area of 12 × 12 mm^2^ centered on the macula, using a raster scanning protocol with 1024 horizontal × 256 vertical A-scans. The axial resolution was 7.3 μm with a depth range of 4.49 mm in tissue.

Established algorithms^25,26^ were used to process the measured raster scan data by deriving the polarization-diverse OCT intensity^27,28^, local phase retardation (measurement of the birefringence magnitude), and the orientation of the OA of the birefringence. The data from the vertical raster scans were processed to generate the intensity, birefringence, and OA images for each eye. To further improve the image quality, the axial pixel separation was increased, allowing for an enhanced localization of the axially accumulated Jones matrix, with a necessary correction used to resolve the optic axis at each axial depth.^19^ The intrinsic optical anisotropy of the fibrous collagen enables axis orientation mapping of the collagen organization with high contrast. This then provides detailed information of the alignment of the collagen fibers.^29,30^

The scleral collagen fibers have an interwoven arrangement, which is mostly beyond the resolution of the OCT.^31^ Thus, we observed the net birefringence from these fibers, where individual fiber birefringence was partially canceled out. The OA of this net birefringence indicated a preferential orientation of the collagen fibers, which have been studied by wide-angle X-ray scattering, small angle light scattering, second harmonic generation, and polarized light microscopy.^10,32,33^ Unlike these techniques, PS-OCT provides depth-resolved measurements of the net birefringence *in vivo* and the preferential orientation.^34^ Volumetric streamline rendering was performed using the ParaView software (version 5.12.0; Kitware Inc., New York, USA) for visualizing the OA that represents the orientation of fibrous tissue as in the earlier described method.^19^

### General Ophthalmic Examinations

Each patient underwent comprehensive ophthalmic examinations to measure the refractive error (spherical equivalent) and the axial length (AL, IOL Master 700; Carl Zeiss Meditec Co., Jena, Germany), and the ultra-widefield fundus photographs (Optos PLC, Dunfermline, United Kingdom). Based on the fundus photographs, we assessed the presence of myopic maculopathy according to the META-PM classification.^35^

### Statistical Analyses

The descriptive parameters are presented as the means ± standard deviations (SD). The Mann-Whitney U test was used to determine the significance of the differences in continuous variables. The Chi-squared test was used to determine the significance of the differences for categorical variables. All statistical analyses were performed using SPSS version 29.0 (IBM SPSS Inc., Armonk, USA). *P* values <0.05 were accepted to be statistically significant.

## Results

### Demographics of patients

Eighty-eight eyes of 60 patients with high myopia whose posterior staphyloma was confirmed in the UWF-OCT images underwent PS-OCT image processing. Thirty-six eyes of 27 patients (7 men, 20 women) met the inclusion criteria and were further analyzed. Their mean age was 67.1 ± 10.7 years. The mean AL was 28.75 ± 2.11 mm, and the mean refractive error (spherical equivalent) of 17 eyes was −10.92 ± 4.87 D, excluding 10 eyes with an intraocular lens (IOL; Table 1). Twenty-eight eyes (77.8%) of 22 patients had a macular staphyloma, and eight eyes (22.2%) of 5 patients had an inferior staphyloma (Table 1). The clinical and ocular characteristics of these 2 types of staphylomas were summarized in Table 2. No statistically significant differences were detected in the evaluated parameters between the two groups.

**Table 1:** Clinical characteristics of highly myopic eyes with posterior staphyloma.

| Parameters | Values |
| --- | --- |
| Numbers of eyes (patients) | 36 (27) |
| Gender |  |
| Female | 20 |
| Male | 7 |
| Age, years (mean ± SD, range) | 67.08 ± 10.69 (38 to 87) |
| Axial length, mm (mean ± SD, range) | 28.75 ± 2.11 (26.84 to 33.55) |
| Refractive error, D (mean ± SD, range) | -10.92 ± 4.87 (-6.75 to -20.00) |
| Types of posterior staphyloma | (except for 10 eyes with IOL) |
| Inferior staphyloma, no. of eyes |  |
| Macular staphyloma, no. of eyes | 8 (22.2%) |
|  | 28 (77.8%) |

**Table 2:** Clinical features of eyes with different types of posterior staphylomas.

| Parameters | Inferior Staphyloma | Macular Staphyloma | *P*-value |
| --- | --- | --- | --- |
| Numbers of eyes (patients) | 8 (5) | 28 (22) | NA |
| Gender |  |  | 0.173<sup>†</sup> |
| Female | 4 | 16 |  |
| Male | 1 | 6 |  |
| Age, years (mean ± SD, range) | 64.79 ± 3.09 (38 to 79) | 67.92 ± 1.69 (51 to 87) | 0.161<sup>*</sup> |
| Axial length, mm (mean ± SD, range) | 27.78 ± 1.33 (26.84 to 29.06) | 29.48 ± 1.31 (28.76 to 33.55) | 0.079<sup>*</sup> |
| Refractive error, D (mean ± SD, range) | -7.45 ± 2.55 (-6.75 to -10.63)<br>(except for 2 eyes with IOL) | -9.71 ± 2.85 (-7.88 to -20.00)<br>(except for 8 eyes with IOL) | 0.121<sup>*</sup> |
| Category of myopic maculopathy<sup>‡</sup><br>(Numbers of eyes) |  |  | 0.771<sup>†</sup> |
| C2 (diffuse atrophy) | 6 | 11 |  |
| C3 (patchy atrophy) | 2 | 13 |  |
| C4 (macular atrophy) | 0 | 4 | 0.372<sup>†</sup> |
| Types of staphyloma edge |  |  |  |
| Upper edge, no. of edges | 8 | 19 |  |
| Lower edge, no. of edges | 0 | 11 |  |
| Total, no. of edges | 8 | 30 |  |

### PS-OCT Images of Macular Staphylomas

In the 28 eyes with a macular staphyloma, 30 edges were observed in the scanned area. The upper edge was detected in 17 eyes, the lower edge in 9 eyes, and both edges were observed in the remaining 2 eyes. Then, 19 upper and 11 lower edges were examined in detail. No other edges, such as the temporal edge (temporal to the macula) were observed in these 30 eyes.

#### Upper Edges of Macular Staphylomas

In the PS-OCT OA images of the 19 upper edges of macular staphylomas, 3 (15.8%) had a gathering of mainly horizontal fibers (Figures 1D & 2D), 13 (68.4%) had a thickening and protrusion of a mixture of horizontal and oblique fibers (Figure 3D), and 2 (10.5%) had an aggregation of oblique fibers alone. Only one edge had a thickening of the vertical fibers. Streamlined images were successfully reconstructed in 8 eyes; and an arc-like gathering of the horizontal fibers was observed along the upper edge of staphylomas (Figure 3G). Oblique fibers oriented in opposite directions were observed above and below this edge as if they were dragged toward the upper edge. The birefringence images showed a mixture of high and low birefringence at 11 edges (57.9%; Figures 1E, 3E), and relatively high birefringence at 8 edges (42.1%; Figure 2E).

**Figure 1.**
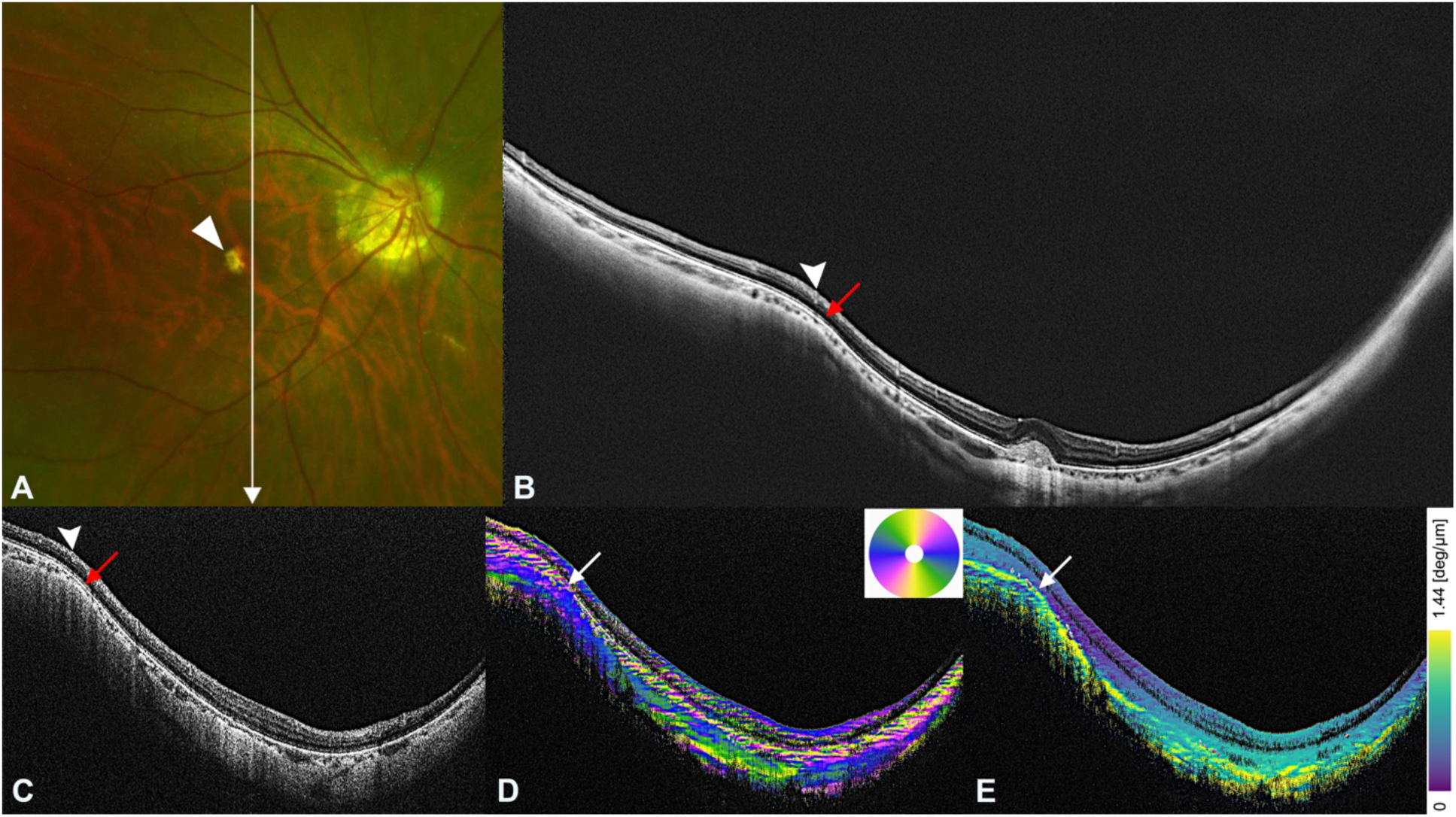
Polarization-sensitive OCT images of the upper edge of an eye with wide macular staphyloma. **A**. Right fundus photograph of a 51-55-year-old woman with a refractive error of −8.5 diopters and an axial length of 27.92 mm showing a scarred macular neovascularization (MNV) and a small atrophy (arrowhead) temporal to the MNV. The arrow points to the PS-OCT scan line in images C-E. **B.** A vertical ultra-widefield optical coherence tomography (UWF-OCT) image showing the staphyloma edge superior to the macula (arrow). A macular MNV is also seen. A retinal vein (arrowhead) is situated over the staphyloma edge. **C.** A vertically scanned PS-OCT intensity image nasal to the fovea shows a steep staphyloma edge superior to the macula (arrow). Above the staphyloma edge, a retinal vein (arrowhead) can be seen. **D.** In a vertical scan of the optic axis image indicates an aggregation of horizontally oriented blue fibers at the staphyloma edge (arrow). **E.** Vertical section of the birefringence image showing a mixture of high (yellow) and low (green) birefringence at the staphyloma edge (arrow). deg = degrees.

**Figure 2.**
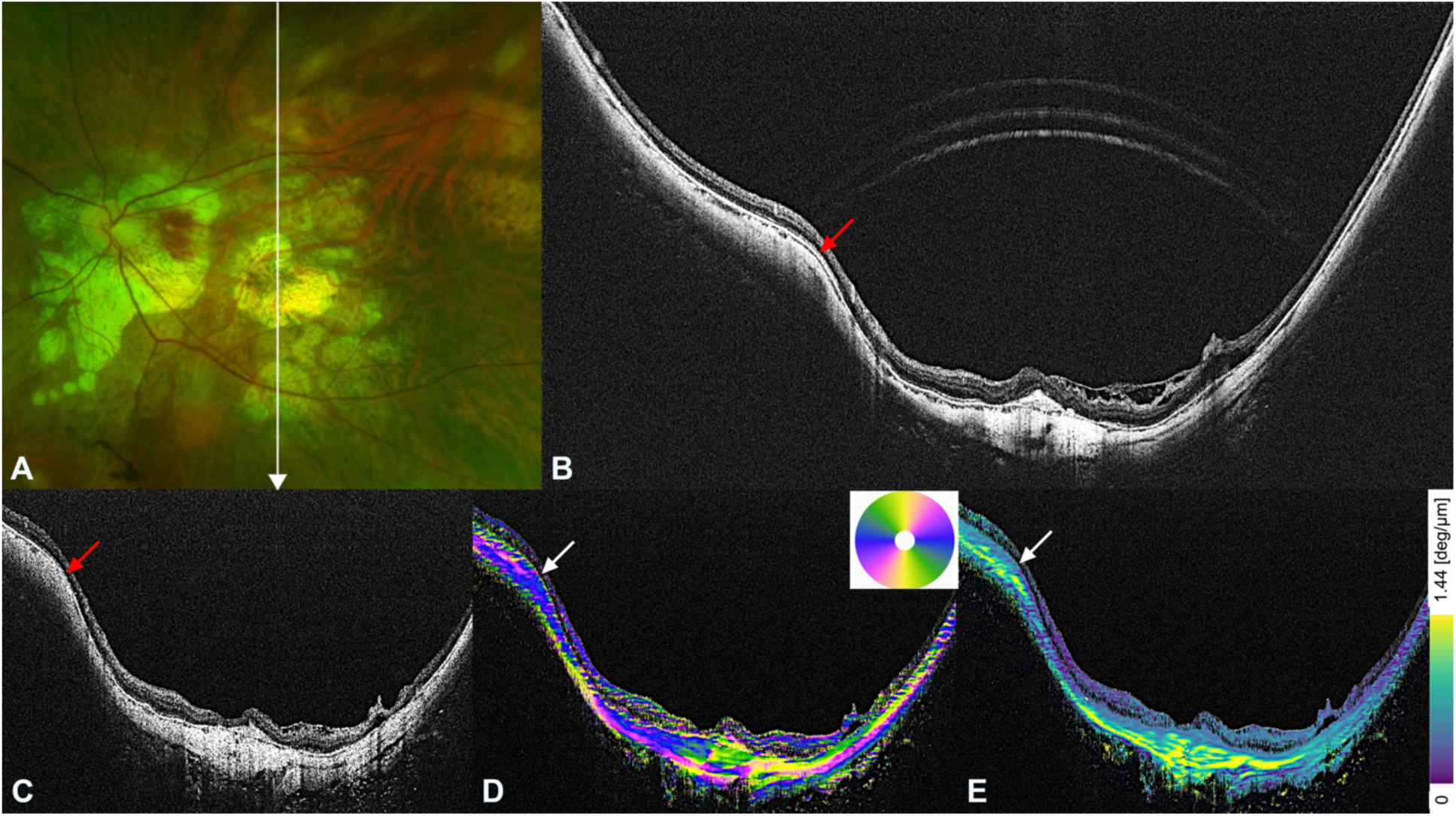
Polarization-sensitive OCT images of the upper edge of an eye with wide macular staphyloma. **A.** Left fundus photograph of a 71-75-year-old woman with an implanted intraocular lens and an axial length of 30.0 mm showing macular atrophy and a wide-ranging patchy choroidal atrophy inferior to the macular. The arrow indicates the PS-OCT scan line in images C-E. **B.** A vertical ultra-widefield optical coherence tomography (UWF-OCT) image shows the upper staphyloma edge superior to the macula (arrow). **C.** A vertically scanned PS-OCT intensity image shows a staphyloma edge superior to the macula (arrow). **D.** In the vertical scan of the optic axis image, horizontally running blue fibers are densely gathered at the staphyloma edge (arrow). **E.** Vertical section of the birefringence image showing relatively high birefringence fibers (yellow) at the staphyloma edge (arrow). deg = degrees.

**Figure 3.**
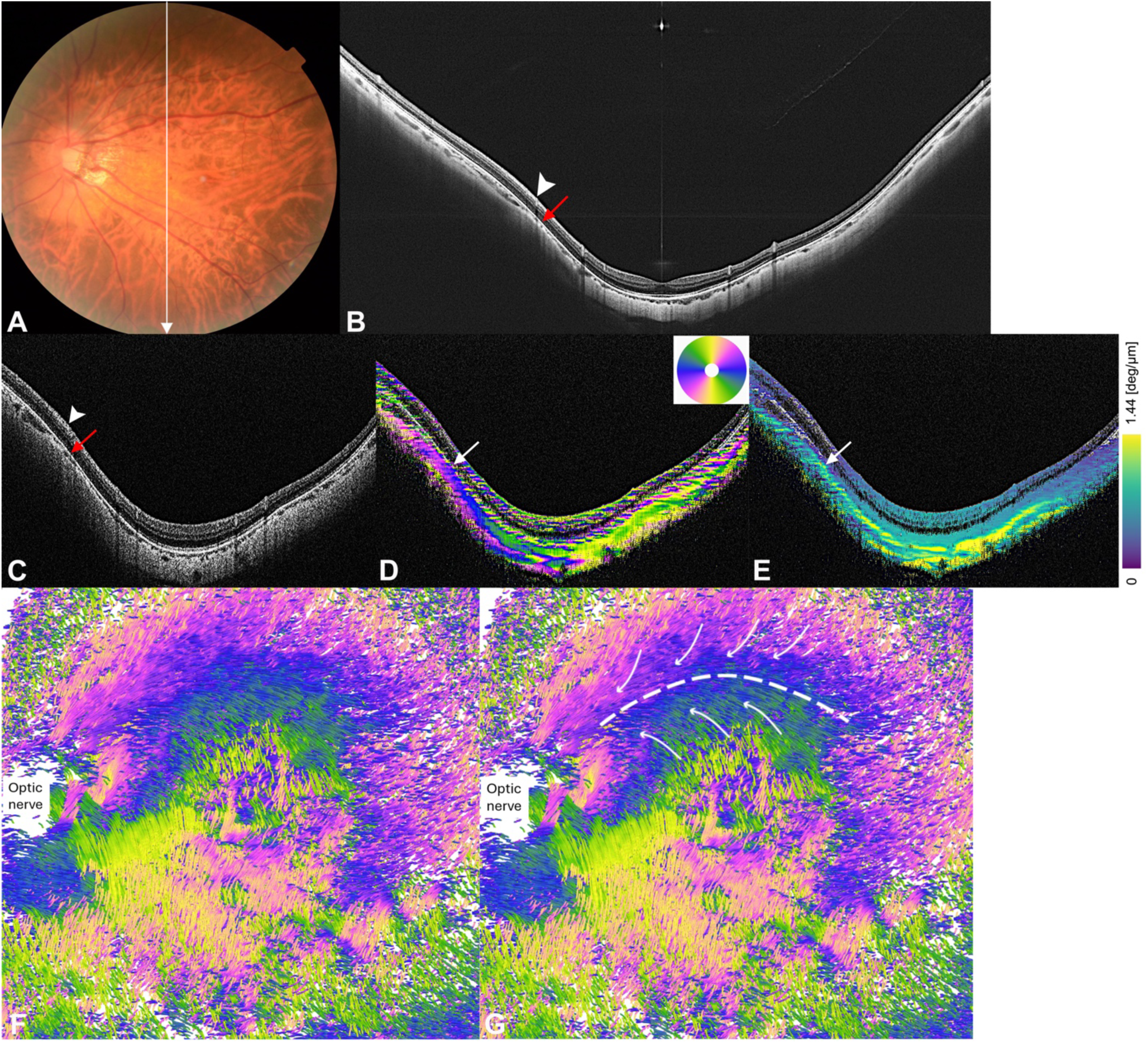
Polarization-sensitive OCT images of the upper edge of an eye with narrow macular staphyloma. **A.** Left fundus photograph of a 56-60-year-old woman with a refractive error of −12.3 diopters and an axial length of 27.56 mm showing diffuse choroidal atrophy. The arrow points to the PS-OCT scan line in images C-E. **B.** A vertical scan of ultra-widefield optical coherence tomography (UWF-OCT) image showing a mild staphyloma edge superior to the macula (arrow). A retinal vein (arrowhead) is situated above the staphyloma edge. **C.** A vertically scanned PS-OCT intensity image across the macula shows a mild staphyloma edge superior to the macula (arrow), with a retinal vein (arrowhead) located over the staphyloma edge. **D.** A vertical scan of the optic axis image shows the thickening of fibers in a mixture of oblique (pink) and horizontal (blue) orientations at the staphyloma edge (arrow). **E.** Vertical section of the birefringence image showing a mixture of high (yellow) and low (green) birefringence at the staphyloma edge (arrow). **F and G.** *En face* streamlined images of the sclera fibers**. (F).** A streamlined image rendered by ParaView viewed from inside the eye. **(G).** An arc-like aggregation of inner horizontal scleral fibers (blue) is observed along the staphyloma edge (white dotted curved line). The orientation of scleral fibers is in the opposite direction above and below the staphyloma edge (curved arrows). deg = degrees.

#### Lower Edges of Macular Staphyloma

In the PS-OCT OA images of the 11 lower edges of macular staphylomas, 5 (45.4%) had an aggregation of a mixture of horizontal and oblique fibers, 2 (18.2%) had a gathering of mainly horizontal fibers, and 4 (36.4%) had a thickening of vertical fibers (Figure 4D and Figure 5D). Although the number of eyes examined was small, the aggregation of vertical fibers was more common at the lower edges than at the upper edges (36.4% vs. 5.3%). The birefringence images showed a mixture of high and low birefringence at 6 edges (54.5%) and relatively high birefringence at 4 edges (36.4%; Figure 4E and Figure 5E). Only one edge showed a relatively low birefringence.

**Figure 4.**
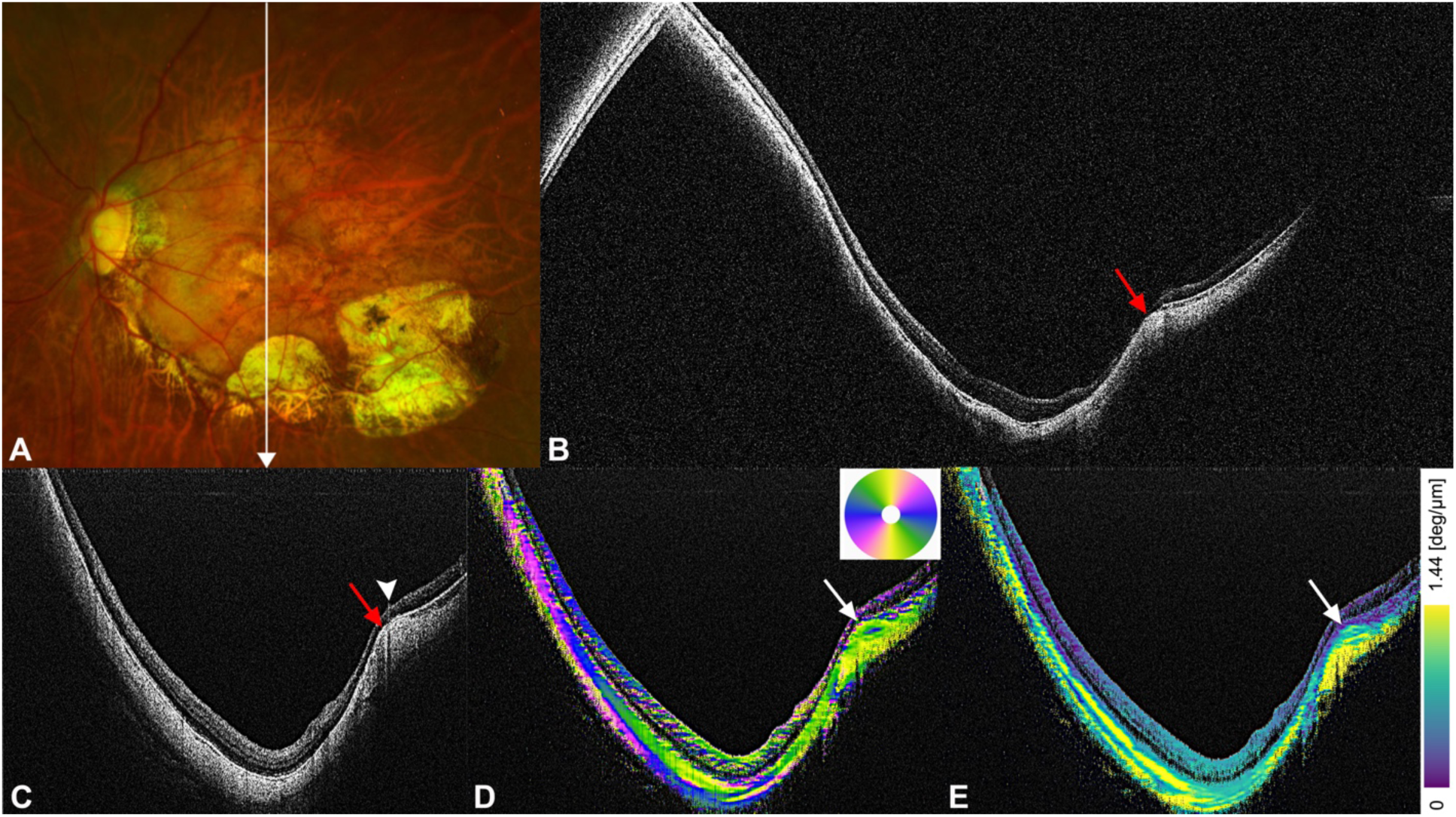
Polarization-sensitive OCT images of the lower edge of an eye with wide macular staphyloma. **A.** Left fundus photograph of a 71-75-year-old woman with an implanted intraocular lens and an axial length of 33.21 mm showing two patchy choroidal atrophies with well-demarcated borders along the lower-temporal vein. The lower edge of staphyloma is seen along this vein. The arrow indicates the PS-OCT scan line in images C-E. **B.** A vertical ultra-widefield optical coherence tomography (UWF-OCT) image showing the staphyloma edge inferior to the macula (arrow). **C.** A vertically scanned PS-OCT intensity image showing a steep staphyloma edge inferior to the macula (arrow). On the staphyloma edge, a retinal vein can be observed (arrowhead). **D.** In the vertical scan of the optic axis image, mainly vertical fibers (yellow) are gathered at the staphyloma edge (arrow). **E.** Vertical section of the birefringence image showing relatively high birefringence fibers (yellow) at the staphyloma edge (arrow). deg = degrees.

**Figure 5.**
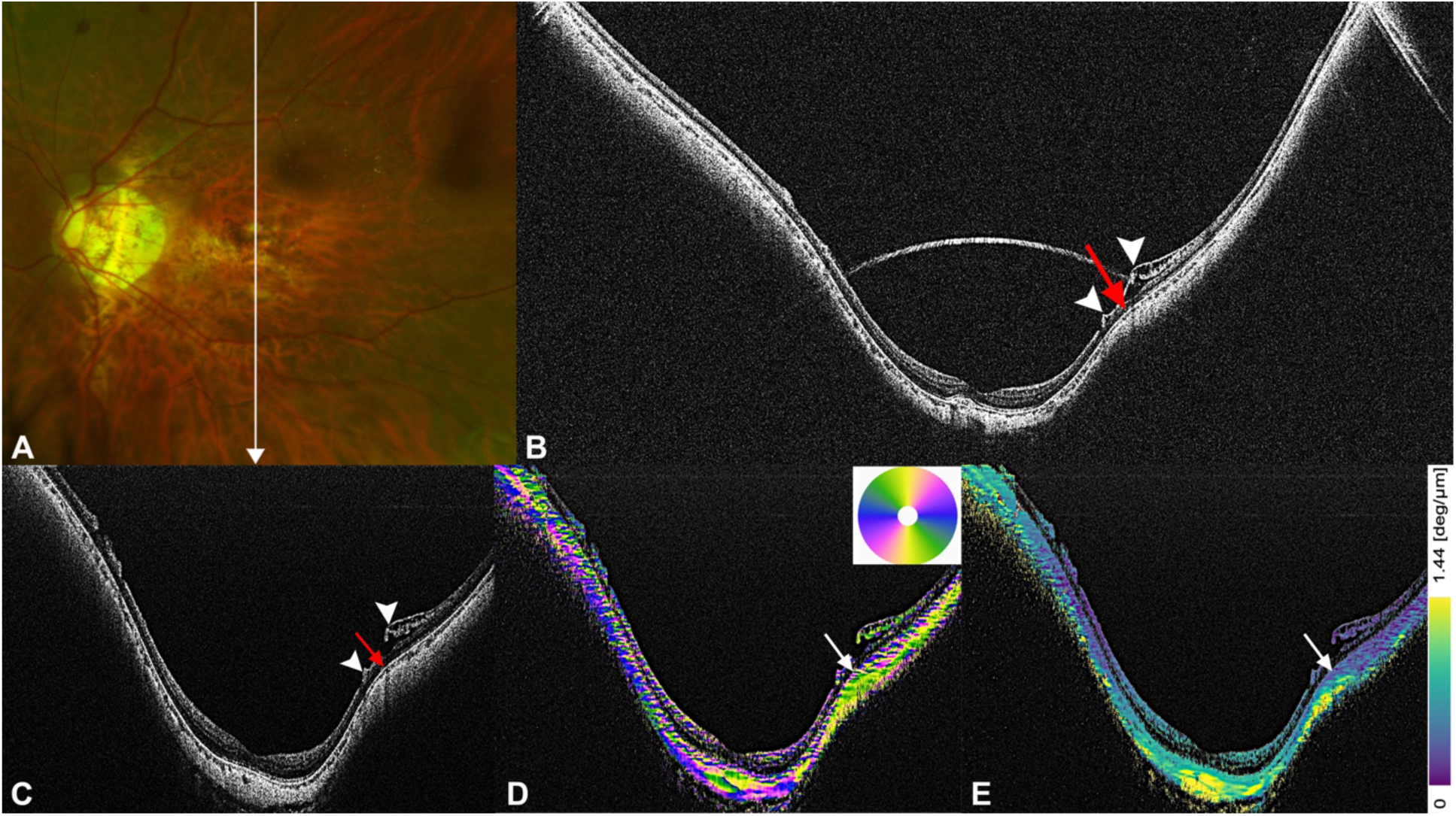
Polarization-sensitive OCT images of the lower edge of an eye with narrow macular staphyloma. **A.** Left fundus photograph of a 51-55-year-old woman with a refractive error of −14.5 diopters and an axial length of 30.76 mm showing diffuse choroidal atrophy. The arrow points to the PS-OCT scan line in images C-E. **B.** A vertical ultra-widefield optical coherence tomography (UWF-OCT) image showing the staphyloma edge inferior to the macula (arrow). Retinal vessels (arrowheads) are seen on the top of the staphyloma edge. An inner and outer retinoschisis surrounding the retinal vessel can also be seen. **C.** A vertically scanned PS-OCT intensity image shows the lower staphyloma edge located beneath the retinal vessels (arrowheads). It is accompanied by retinoschisis surrounding these vessels. **D.** In the vertical scan of the optic axis image, vertical fibers are predominantly thickened at the staphyloma edge (arrow). **E.** Vertical section of the birefringence image showing relatively high birefringence fibers (yellow) at the staphyloma edge (arrow). deg = degrees.

### PS-OCT Images of Inferior Staphylomas

In the eight eyes with an inferior staphyloma, only the upper edges across the macula were observed. Eight edges were identified in the scanned areas. The OA images revealed an aggregation of a mixture of horizontal and oblique fibers at 6 edges (75%), and the gathering of horizontal fibers alone at 2 edges (25%, Figure 6E). In the birefringence images, the scleral fibers at the staphyloma edge showed mostly high birefringence (Figure 6F) at five edges (62.5%), and a mixture of high and low birefringence at three edges (37.5%). Streamlined images were successfully reconstructed in four eyes; and a more linear structure was observed along the upper staphyloma edge (Figure 6H). Near the nasal margin of the upper edge, the oblique fibers were observed to be aligned in opposite directions and appeared to merge with this edge.

**Figure 6.**
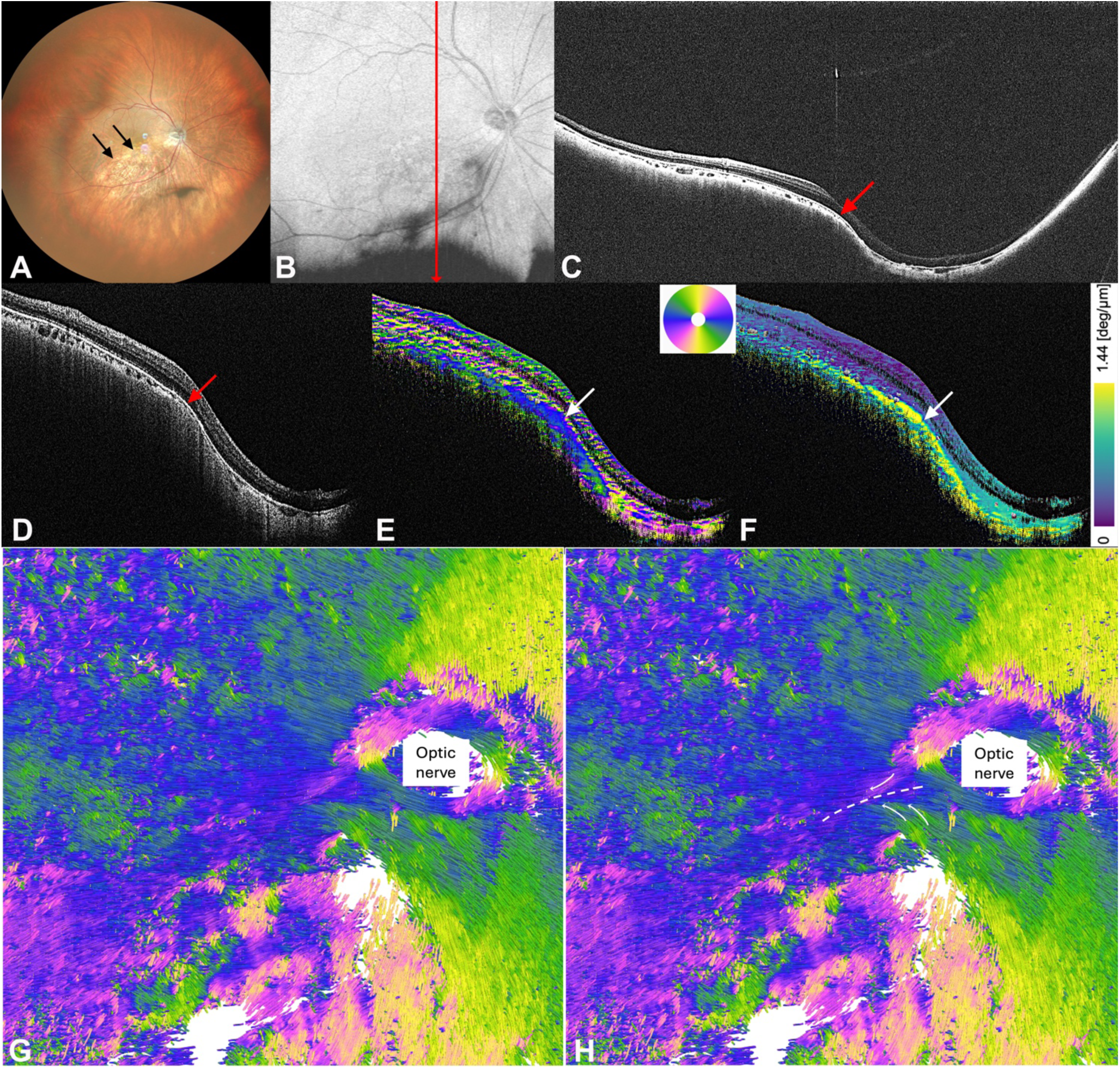
Polarization-sensitive OCT images of the upper edge of an eye with inferior staphyloma. **A.** Right fundus photograph of a 71-75-year-old woman with an implanted intraocular lens and an axial length of 26.88 mm showing the depigmented margin of the inferior staphyloma across the macula (arrows). **B.** The red arrow in the *En face* OCT intensity projection indicates the PS-OCT scan line in images D-F **C**. A vertical scan of ultra-widefield optical coherence tomography (UWF-OCT) image showing the upper edge of a staphyloma extending across the macula (arrow). **D.** A vertically scanned PS-OCT intensity image across the fovea shows the upper edge of the staphyloma (arrow). **E.** A vertical scan of optic axis image showing that the horizontally oriented blue fibers (arrow) are aggregated at the staphyloma edge. **F.** Vertical section of the birefringence image shows high birefringence fibers (yellow, indicated by arrow) at the staphyloma edge. **G and H.** *En face* streamline images of the sclera fibers. **(G).** Interior view of the streamlined image rendered by ParaView. **(H).** A linear structure is observed along the staphyloma edge (indicated by the white dotted line). Near the nasal margin of the staphyloma edge, scleral fibers running in the opposite directions are seen merging into this upper edge from above and below (curved arrows). deg = degrees.

### Retinal and Retinal Vascular Changes at Edges of Macular Staphylomas

Unlike the upper edge of inferior staphylomas, the upper and lower edges of the macular staphylomas were observed near the retinal vascular arcade. We found that major retinal vessels were present at the top of the staphyloma edges in 17 of the 30 edges (56.7%) of the macular staphyloma, 70.6% of the upper edges (Figure 7B), and 45.4% of the lower edges (Figure 5C). Paravascular patchy atrophy was found in 4 of the 17 edges (23.5%) that had retinal vessels over the top of the edge. All four edges were lower edges. In the 17 edges that had retinal vessels over the top, various vascular and paravascular findings were observed: paravascular retinoschisis at one edge (5.9%, Figure 5C), paravascular retinal thinning in 5 edges (29.4%, Figures 7B & 7D) and anterior protrusion of retinal vessels (retinal vascular microfolds) in 5 edges (29.4%, Figures 7D & 7F).

**Figure 7.**
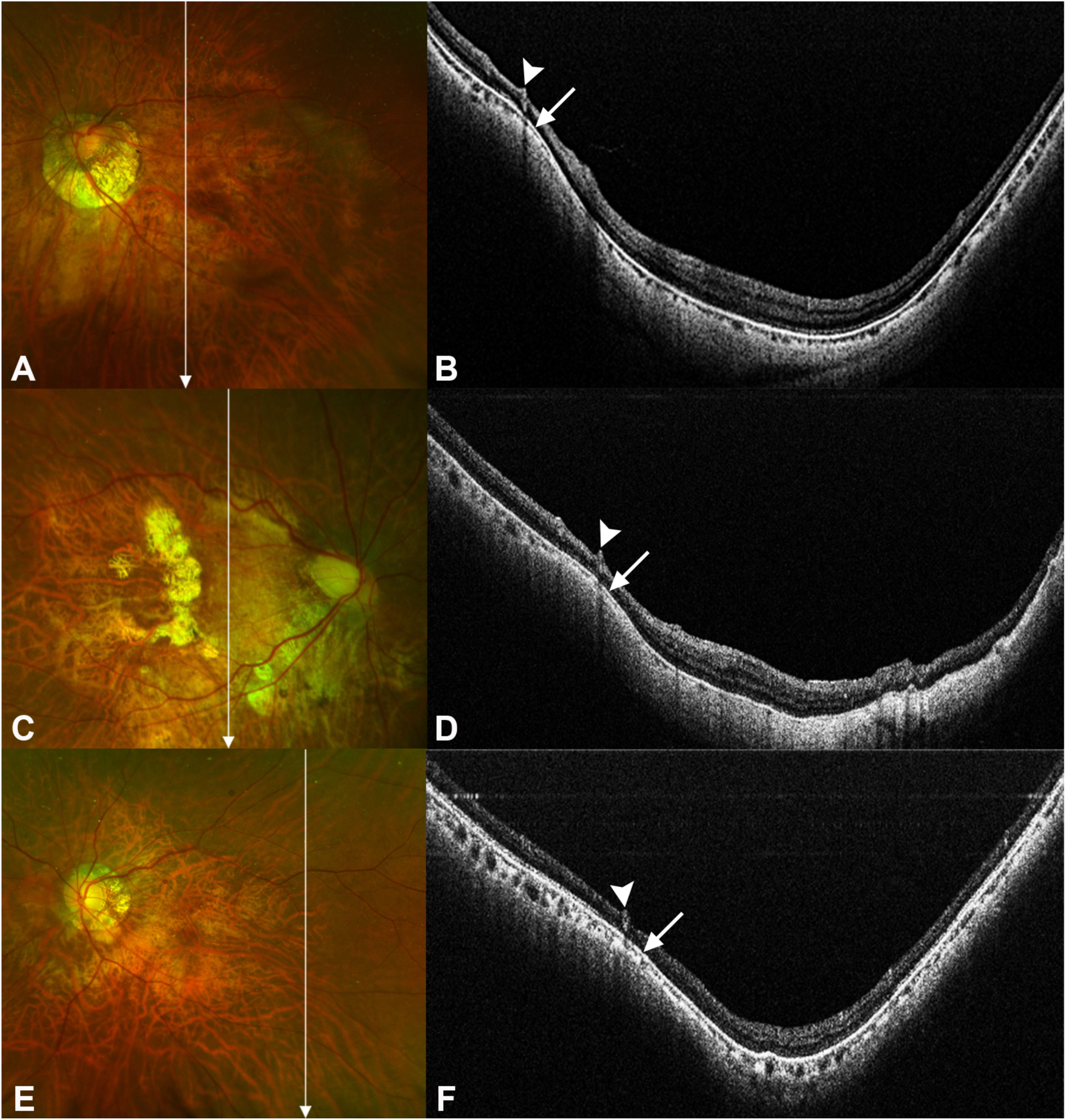
Images of retinal and retinal vascular abnormalities at the edges of macular staphyloma. **A.** Fundus photograph of the left eye of a 66-70-year-old woman with an axial length of 30.03 mm showing mild diffuse choroidal atrophy. Arrow indicates the PS-OCT scan line in image B. **B.** Vertically scanned PS-OCT intensity image shows the retinal vein (arrowhead) located over the upper staphyloma edge (arrow). Retina thinning around this vessel is also noted. **C.** Fundus photograph of the right eye a 71-75-year-old woman with an axial length of 31.65 mm showing macular neovascularization and macular atrophy. Arrow indicates the PS-OCT scan line in image **D.** A vertically scanned PS-OCT intensity image shows an anterior protrusion of a retinal vessel (arrowhead) over the edge of the staphyloma (arrow), accompanied by thinning of the surrounding retina. **E** Fundus photograph of the left eye of a 51-55-year-old woman with an axial length of 29.92 mm showing diffuse choroidal atrophy. Arrow indicates the PS-OCT scan line in image F. **F.** Vertically scanned PS-OCT intensity image shows an anteriorly protruded retinal vessel (arrowhead) over the staphyloma edge (arrow).

## Discussion

Building on the earlier study using PS-OCT, the OA image of scleral fibers can clearly differentiate the inner and outer layer of sclera in highly myopic eyes: the inner sclera generally shows a radial orientation from the optic nerve to the periphery; the outer sclera shows a vertical orientation in the posterior fundus.^19^ In the current study, we utilized PS-OCT to examine the scleral collagen fibers at the edges of posterior staphylomas in highly myopic eyes in vivo. Our findings showed that 25 (18 upper and 7 lower) of the 30 macular staphyloma edges (83.3%) had an aggregation of inner scleral fibers. Streamlined images revealed an arc-like aggregation of the inner scleral horizontal fibers along the staphyloma edge. Eight upper edges of inferior staphylomas were examined; all had inner scleral fiber aggregation. Streamlined images revealed the linear aggregation of the inner scleral horizontal fibers ran over the macula. Despite the differences in the location of the upper edges between the macular and inferior staphylomas, the PS-OCT findings for staphyloma edges were consistent.

In eyes with staphylomas, the upper edges were typically the most prominent, especially in adults.^36^ The upper edges of macular and inferior staphylomas consisted primarily of aggregated inner scleral fibers similar to those in eyes with DSM. Ohno-Matsui et al.^19^ reported that the outer sclera was not thickened but compressed by the thickened inner sclera. These similarities suggested that the staphyloma edges and DSM may result from an uneven distribution and displacement of the inner scleral fibers. The inner sclera may play a more critical role in the changes of scleral curvature than the outer sclera with each layer contributing differently to maintaining the integrity of the scleral structure. Saito et al.^37^ investigated 511 highly myopic eyes with DSM using UWF-OCT and observed that the eyes with a DSM either lacked staphylomas or presented mostly with wide staphylomas. These findings suggested that the uneven aggregation of the inner scleral fibers may not occur in proximity simultaneously such as macula and vascular arcade.

Conversely, the remaining five macular staphyloma edges consisted primarily of vertical fibers in the outer sclera with four being at the lower edges. The outer scleral fiber aggregation was found more frequent at the lower edges (4 of the 11 lower edges; 36.4%) than in the upper edges (1 of the 19 edges; 5.3%). All four lower edges with outer scleral fiber aggregation were accompanied by an overlying paravascular patchy atrophy (Figure 4D). Due to the limited PS-OCT scan size, only a small number of lower edges were included. Although further studies are required, it is possible that the disappearance of the choroid within the area of paravascular patchy atrophy caused an extreme thinning of the inner sclera leading to a compensatory gathering of the outer sclera. The arrangement of the scleral fibers may vary with the location or degree of scleral protrusion at the edges.

Two of the four edges studied were associated with Bruch’s membrane (BM) defects. A previous study^38^ proposed that BM defects surrounding a DSM result from a focal relaxation of the posterior sclera and insufficient internal pressure from an expanding BM, causing a partial inward sclera bulge. This mechanism might have also been applied to the staphyloma edge formation in these four cases of paravascular patchy atrophy. Future studies with broader scanned areas and more cases, especially those with lower staphyloma edges, are needed to determine the causative factors.

Consistent with a previous study birefringence results of the upper and lower staphyloma edges showed different patterns including a mixture of high and low, and relatively high.^18^ Histological studies indicated that scleral collagen bundles in eyes with posterior staphyloma shift from an interwoven to a lamellar arrangement.^39^ This shift may explain the increased birefringence observed at some staphyloma edges that potentially reduce the resistance to the elongation forces under normal intraocular pressures. Such a weakening could create a region prone to sclera protrusion.^40^ Further biomechanical research is needed to verify this suggestion.

Scleral remodeling of the posterior sclera is a key process in the development of myopia, characterized by a loss of collagen tissue, a thinning of collagen fiber bundles, and a reduction in collagen fibril diameter.^9–11^ These alterations collectively contribute to overall scleral thinning, which is pronounced at staphylomatous areas.^12^ Our findings revealed that the structural changes at the staphyloma edge are not merely a consequence of uniform scleral thinning; rather, they show an uneven aggregation of collagen fibers arranged in a more aligned pattern. This localized aggregation may exacerbate scleral bulging in highly myopic eyes, where reduced scleral stiffness renders the tissue more susceptible to deformation under intraocular pressure (IOP), ultimately leading to staphyloma formation. Braeu et al.^41^ proposed a three-dimensional finite element model suggesting that a local scleral weakening could be the sole trigger for the formation and development of posterior staphylomas under the normal IOP. This synergy between pathological evidence and mathematical model supports the perspective that targeted reinforcement of the scleral weak points may hold therapeutic potential for staphylomas.

In addition to the PS-OCT findings, we noted a notable colocalization of the major retinal vessels with the upper/lower staphyloma edges. Major retinal vessels were located at the top of 17 of the 30 staphyloma edges (56.7%). Various vascular and paravascular abnormalities were also identified, including retinal vascular microfolds, retinal thinning around protruded retinal vessels, and paravascular retinoschisis. Whether these abnormalities contribute to the development of the staphyloma edge or are coincidental was not determined. Notably, the upper and lower edges generally align with the retinal vascular arcades. Takahashi et al.^42^ demonstrated a firm vitreal traction of the retinal vessels in eyes with pathological myopia. This finding suggested that this traction may play a more critical role in the development of myopic macular retinoschisis than foveal traction. This intense vitreous traction along with the retinal vascular stiffness may help prevent a bowing of the sclera posteriorly and thus maintain the anatomical position of the sclera. In normal eyes, the retinal vessels run on the retinal surface and thus maintains a distance from the sclera. However, in pathological myopia, especially in areas of paravascular patchy atrophy, the retina beneath and around the vessels is extremely thin, with vessels sometimes resting on the sclera.^43,44^ Further studies are needed to determine the mechanisms underlying these observations, with a focus on the role of vascular structure and vitreous traction in the formations of staphylomas.

Currently, no effective therapies exist to halt the development or progression of posterior staphylomas. Scleral collagen crosslinking has been reported to prevent axial myopia in animal models by enhancing scleral rigidity;^45,46^ however, its effect on posterior staphylomas remains unclear. Our study may suggest that modulating the architectural organization and biomechanical alignment of scleral collagen fibers may represent a potential therapeutic strategy. Chen et al.^37^ recently reported that mild staphyloma edges in children and adolescents with myopia developed much earlier than previously believed. Longitudinal studies in this population could illuminate when the scleral fiber changes occur before and at the onset of staphyloma edge formation.

There are several limitations to this study. First, this study focused exclusively on patients with high myopia limiting the generalization of the findings to individuals with non-high myopia but with posterior staphylomas. Second, this study was descriptive and based on a small number of cases. A larger cohort and quantitative analyses are essential to validate and expend these observations. Third, the lack of long-term follow-up prevented an assessment of longitudinal changes, highlighting the need for future longitudinal investigations to gain a more comprehensive understanding of posterior staphyloma edges. Finally, the study did not evaluate nasal and temporal staphyloma edges because of the vertical scanning protocol and the limited scan width of PS-OCT, resulting in incomplete data of these variants.

In conclusion, the results indicate that the staphyloma edges are predominately comprised of an uneven aggregation of inner scleral fibers with the outer sclera protruding only into areas lacking the inner sclera. Investigating the occurrence and mechanism causing this inner scleral fiber aggregation could provide clues for developing early therapeutic interventions for posterior staphylomas. Additionally, the unusual co-location of the retinal vascular arcade with staphyloma edges may disclose preferential locations for staphyloma formation.

## Data Availability

All data produced in the present study are available upon reasonable request to the authors

## Acknowledgements

The authors thank Professor Emeritus Duco Hamasaki of the Bascom Palmer Eye Institute at the University of Miami in Miami, Florida, for his critical discussions and thorough editing of the manuscript.

## Notes

**Financial Support:** This study was supported by JST SPRING, Grant Number JPMJSP2120 (Yijin, Wu) and received partial research funding from Tomey Corporation. Tomey Corporation had no role in the design and conduct of this research.

**Conflict of Interest Disclosures:** M.Y. and M.O. are employees of Tomey Corporation. Dr Ohno-Matsui reported receiving a research grant from Tomey Corporation and being a consultant for Santen and CooperVision outside the submitted work. Dr Yamanari reported having patents for JP 6463051 B2, US 9593936 B2, EP 2995245 B1, JP 6542178 B2, and JP 7332131 B2 issued to Tomey Corporation. The remaining authors declare no conflicts of interest.

### Competing Interest Statement

M.Y. and M.O. are employees of Tomey Corporation. Dr Ohno-Matsui reported receiving a research grant from Tomey Corporation and being a consultant for Santen and CooperVision outside the submitted work. Dr Yamanari reported having patents for JP 6463051 B2, US 9593936 B2, EP 2995245 B1, JP 6542178 B2, and JP 7332131 B2 issued to Tomey Corporation. The remaining authors declare no conflicts of interest.

### Funding Statement

This study was supported by JST SPRING, Grant Number JPMJSP2120 and received partial research funding from Tomey Corporation. Tomey Corporation had no role in the design and conduct of this research.

### Author Declarations

Ethics Committee of the Institute of Science Tokyo gave ethical approval for this work

